# Genetic analysis of 104 pregnancy phenotypes in 39,194 Chinese women

**DOI:** 10.1101/2023.11.23.23298979

**Authors:** Han Xiao, Linxuan Li, Meng Yang, Jingyu Zeng, Jieqiong Zhou, Ye Tao, Yan Zhou, Mingzhi Cai, Jiuying Liu, Yushan Huang, Yuanyuan Zhong, Panhong Liu, Zhongqiang Cao, Hong Mei, Xiaonan Cai, Liqin Hu, Rui Zhou, Xun Xu, Huanming Yang, Jian Wang, Huanhuan Zhu, Aifen Zhou, Xin Jin

**Affiliations:** Institute of Maternal and Child Health, Wuhan Children’s Hospital (Wuhan Maternal and Child Health care Hospital), Tongji Medical College, Huazhong University of Science and Technology, Wuhan, China; BGI-Shenzhen, Shenzhen, China; College of Life Sciences, University of Chinese Academy of Sciences, Beijing 100049, China; College of Life Sciences, Northwest A&F University, Yangling, Shaanxi, China; Department of Obstetrics, Wuhan Children’s Hospital (Wuhan Maternal and Child Health care Hospital), Tongji Medical College, Huazhong University of Science and Technology, Wuhan, China; BGI-Wuhan Clinical Laboratories, BGI-Shenzhen, Wuhan, China; Guangdong Provincial Key Laboratory of Genome Read and Write, Shenzhen, China; Guangdong Provincial Academician Workstation of BGI Synthetic Genomics, Shenzhen, China; James D. Watson Institute of Genome Sciences, Hangzhou, China; The Cancer Hospital of the University of Chinese Academy of Sciences (Zhejiang Cancer Hospital), Institute of Basic Medicine and Cancer (IBMC), Chinese Academy of Sciences, 16 Hangzhou, China; BGI, BGI-Shenzhen, Shenzhen, China; School of Medicine, South China University of Technology, Guangzhou, China

## Abstract

Maternity is a special period in a woman’s life that involves substantial physiological, psychological, and hormonal changes. These changes may cause alterations in many clinical measurements during pregnancy, which can be used to monitor and diagnose maternal disorders and adverse postnatal outcomes. Exploring the genetic background of these phenotypes is key to elucidating the pathogenesis of pregnancy disorders. In this study, we conducted a large-scale molecular biology analysis of 104 pregnancy phenotypes based on genotype data from 39,194 Chinses women. Genome-wide association analysis identified a total of 407 trait-locus associations, of which 75.18% were previously reported. Among the 101 novel associations for 37 phenotypes, some were potentially pregnancy-specific and worth further experimental investigation. For example, *ESR1* with fasting glucose, hemoglobin, hematocrit, and several leukocytoses; *ZSCAN31* with blood urea nitrogen. We further performed pathway-based analysis and uncovered at least one significant pathway for 24 traits, in addition to previously known functional pathways, novel findings included birthweight with “Reactome signaling by NODAL”, twin pregnancy with “Reactome mitotic G1-G1/S phases”. The partitioning heritability analysis recapitulated known trait-relevant tissue/cell types, and also discovered interesting results including twin pregnancy with “embryoid bodies” cell-type enrichment, the delivery type cesarean section with “fallopian tube”, and birth weight with “ovary and embryonic stem cells”. In terms of both sample size and the variety of phenotypes, our work is one of the largest genetic studies of pregnancy phenotypes across all populations. We believe that this study will provide a valuable resource for exploring the genetic background of pregnancy phenotypes and also for further research on pregnancy-related diseases and adverse neonatal outcomes.

## Introduction

The clinical phenotypes (e.g., serum and urinary test results) are commonly used for disease detection and diagnosis. Understanding their genetic architecture is key to elucidating disease etiology. Up until now, many genome-wide association studies (GWAS) have been performed to explore the genetic background of these phenotypes, including hematological ^1,2^, liver-related^3–5^, kidney-related^6,7^, metabolic^8,9^, protein^10,11^, and urinary^12,13^. Many biobanks and consortiums also allow large-scale genetic analysis for a vast amount of clinical laboratory measurements, such as UK Biobank^14^, BioBank Japan Project (BBJ)^11^, FinnGen biobank^15^, and DIAGRAM consortium^14^.

The serum and urinary test results during pregnancy are critical for assessing maternal and fetal health status and predicting adverse postnatal outcomes^16–18^. For example, the high maternal glucose level is an indication of the risk of developing gestational diabetes^19^, and abnormal maternal hemoglobin levels may be a warning of preterm birth^20,21^. However, there are few studies investigating the genetic background of the laboratory features during pregnancy. The aforementioned studies and biobanks were typically designed for general adults, not the pregnant population. In recent years, non-invasive prenatal testing (NIPT) has become extensively used to provide pregnant women with a sensitive noninvasive screening option for chromosomal disorders of fetuses^22^. This technology detects whether the fetus has the three major chromosomal disorders while generating high-throughput maternal genotype data. Our team previously demonstrated that the NIPT sequencing data could be used for genetic studies, including variant calling, population history, viral infection patterns, and genome-wide association study^23^. We have proved that with highly accurate imputation performance for the low sequencing depth NIPT data, the GWAS analysis could maintain high statistical power in identifying trait-associated genetic variants.

In this study, based on the genotype data from 39,194 pregnant women who underwent the NIPT test, we investigated the genetic background of 104 pregnancy phenotypes, including maternal phenotypes (e.g., women’s BMI, blood pressure), postnatal outcomes (e.g., birthweight, delivery option), and laboratory measurements (e.g., hematological, urinalysis, hormone, infection). The GWAS analysis identified 407 genome-wide significant associations involved with 66 phenotypes. The majority (75.18%) of these trait-locus associations were previously identified in either European or Asian populations, for example, the BMI with *FTO*, the C-reactive protein level with *CRP*, the serum bilirubin levels with the UDP Glucuronosyltransferase Family 1 Members (e.g., *UGT1A6*). In addition to recapitulating known findings, we also discovered 101 novel trait-locus associations for 37 phenotypes, for example, thyroid hormone triiodothyronine (FT3) and *ABO*, urine glucose levels and *CDK12*, and total bile acid and *SLC39A9*. We also filled the gap of no GWAS results for some phenotypes in the GWAS Catalog, such as prealbumin (transthyretin), platelet-large cell ratio, and mucus in urine. Interestingly, we discovered some potentially pregnancy-specific associations, such as *ESR1* with fasting serum glucose, *ESR1* with several types of leukocytosis, *ZSCAN31* with blood urea nitrogen, and *ABCB4* with γ-glutamyl transferase. The functional enrichment and partitioning heritability analysis revealed inspiring results worthy of further exploration. To name one, for birthweight, the identified functional pathway is “Reactome signaling by NODAL” (regulating embryonic development) and the relevant cell types are ovary and embryonic stem cells. Our work is the first one to implement such a large genetic study of pregnancy phenotypes and all of the GWAS summary statistics results are publicly available for other researchers to use. These results will certainly provide a theoretical basis and reference for the study of genetic mechanisms of maternal disorders and postnatal outcomes.

## Results

### Imputation performance of ultra-low coverage WGS data

In our dataset, 39,194 pregnant women took the NIPT test and had genotype data with an average of data volume 476 MB (sequencing depth was approximately 0.15X). We removed samples with sequencing depth less than 0.05X and mapping rate less than 90% and 38,668 samples remained in further analysis. After genotype imputation in STITCH, there were 8,134,302 genotyped SNPs with SNP density provided in Figure 1a. The average imputation accuracy across all SNPs was 81.37% (Figure 1b). In addition, if we focused on only well-imputed SNPs with info-score > 0.4, Hardy-Weinberg equilibrium (HWE) p-value > 1e-6, and minor allele frequency (MAF) > 0.05, the mean imputation accuracy was 90.46% (Figure 1b). The number of samples who took the folate metabolism ability genetic test was 272. The averaged correlation of imputed and true genotypes of three tested variants in genes *MTHFR* and *MTRR* was 0.71 and the maximum value was 0.90 (Figure 1c). These results ensured the high quality and accuracy of imputed genotype data.

**Figure 1.**
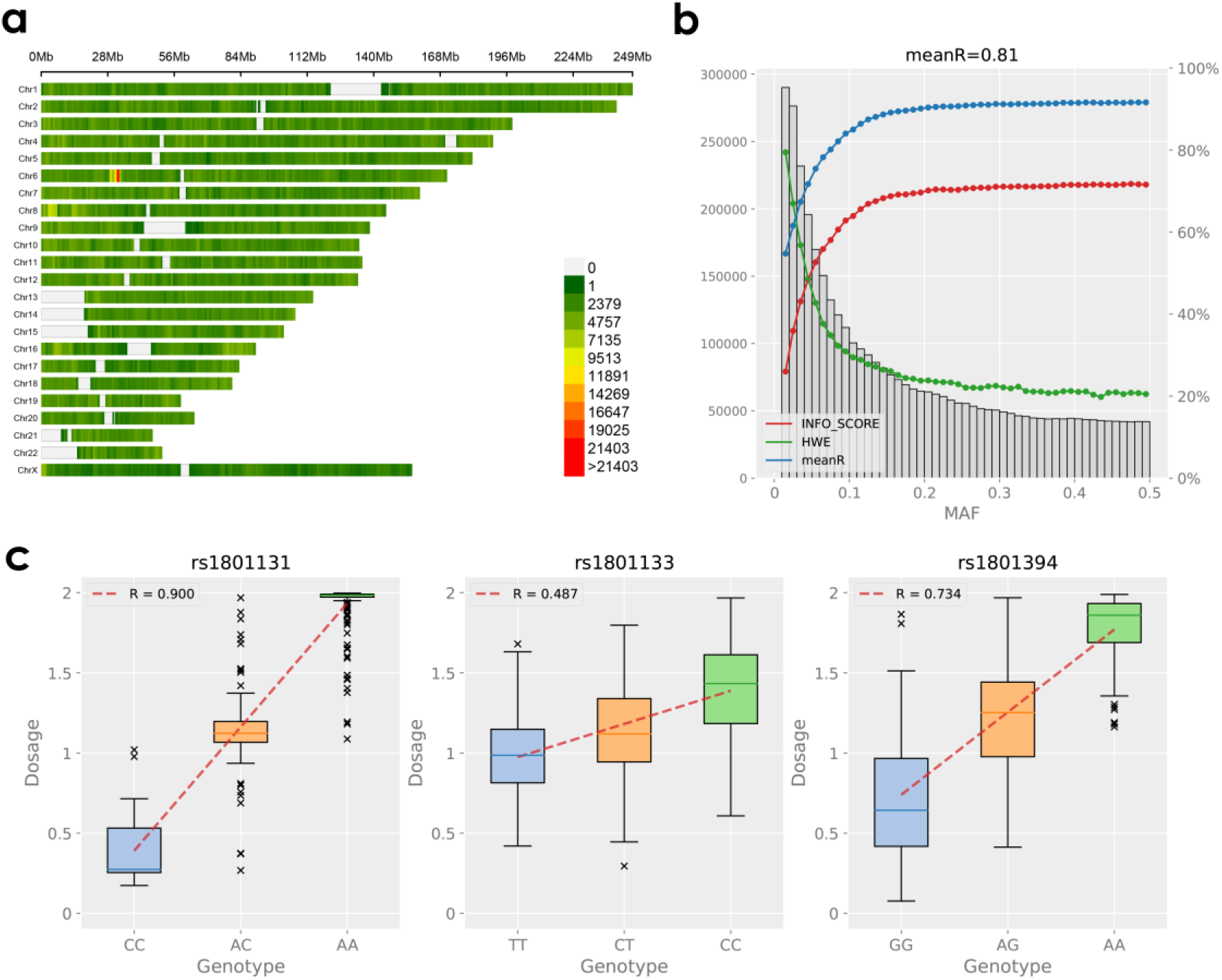
The information on imputed genetic variants. *Notes*: **a)** the SNP density plot of imputed variants, **b)** the genotype imputation accuracy calculated by Pearson’s correlation between high-coverage sequencing data and imputed dosage, and **c)** the boxplot visualization and Pearson’s correlation between imputed and true genotypes of two variants on gene *MTHFR* (rs1801131 and rs1801133) and one variant on gene *MTRR* (rs1801394).

### Genome-wide association analysis of 104 pregnancy traits

We performed GWAS analysis on 104 traits, each with an effective sample size of over 2,000 (Supplementary Figure S1). These traits covered a wide range of clinical measurements, grouped into 11 different categories (Supplementary Figure S1, Table 1): maternal (n=5), postnatal (n=6), electrolyte (n=4), hematological (n=24), hormone (n=4), infection (n=14), kidney-related (n=4), liver-related (n=8), metabolism (n=6), protein (n=5), and urinalysis (n=24). To have a better visualization of trait distribution and pairwise association, we provided a convoluted figure for displaying the frequency of quantitative traits (histogram), the category of binary traits (bar plot), the relationship between two quantitative traits (scatter chart), one quantitative trait and one binary trait (box plot), two binary traits (2*2 contingency table), and the results of statistical inference for testing the corresponding correlations (Supplementary Figure S2). The chromosome-based Circos plot presenting the GWAS results of phenotypes with significant signals was provided in Figure 2a. The genomic inflation factors (λ_gc_) for estimating the amount of GWAS test statistics inflation were presented in bar plots (Figure 2b). The λ_gc_’s of all phenotypes were around 1, meaning no evidence of inflation and the GWAS results were reasonable. The individual Manhattan plots were provided in Supplementary Figure S3. For known and novel loci, we colored blank and red, respectively.

**Figure 2.**
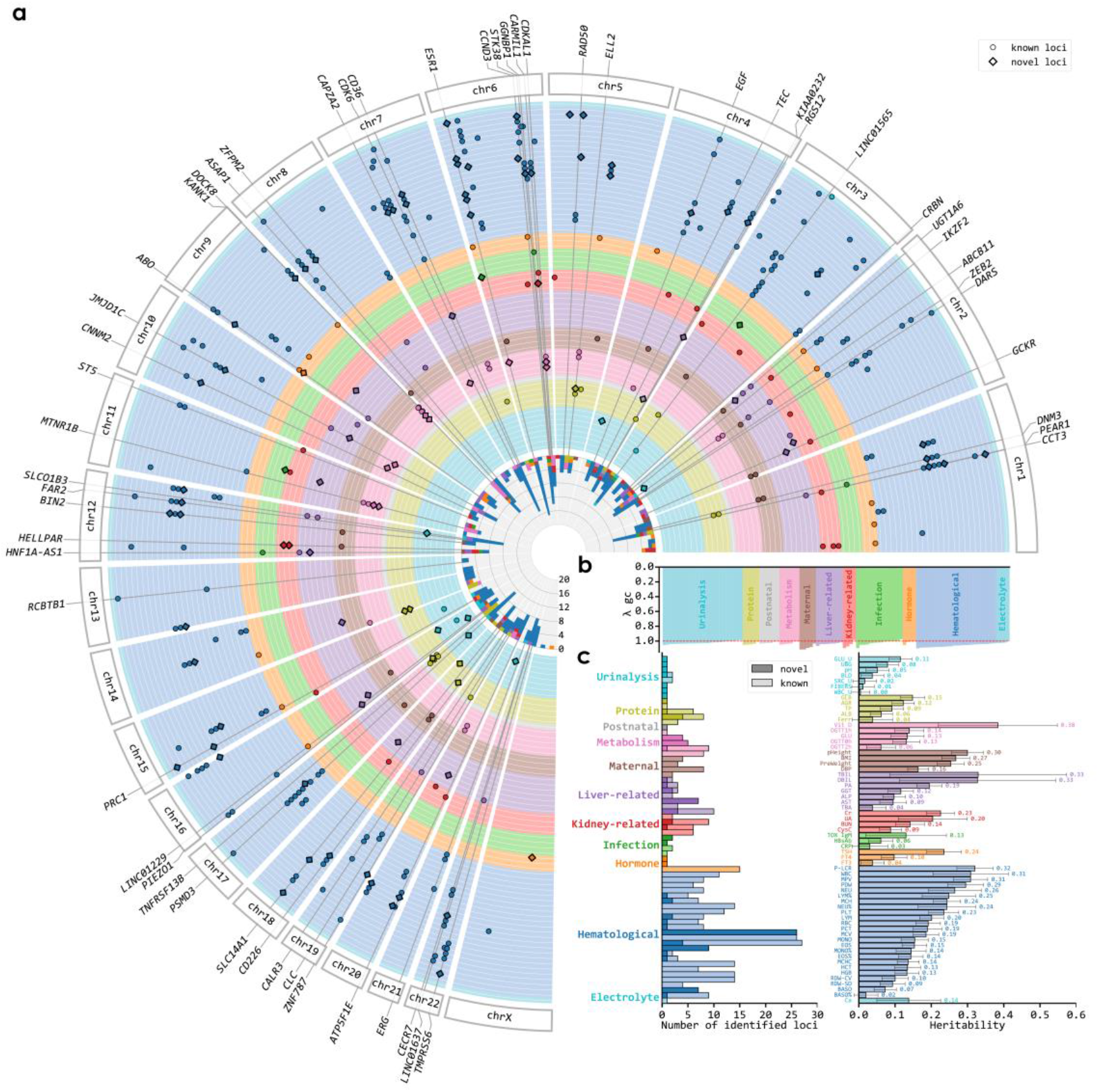
Overview of the identified trait-locus associations. *Notes*: **a)** the circos plot presents the identified trait-locus associations, from inner to outer circle, different colors repretent different phenotype categories, **b)** the genomic inflation factors (λ_gc_) for each phenotype, and **c)** the bar plot (left) provides the number of identified trait-associated loci for each trait, grouped by phenotype categories; and the bar plot (right) provides the heritability value for each phenotype.

**Table 1.**
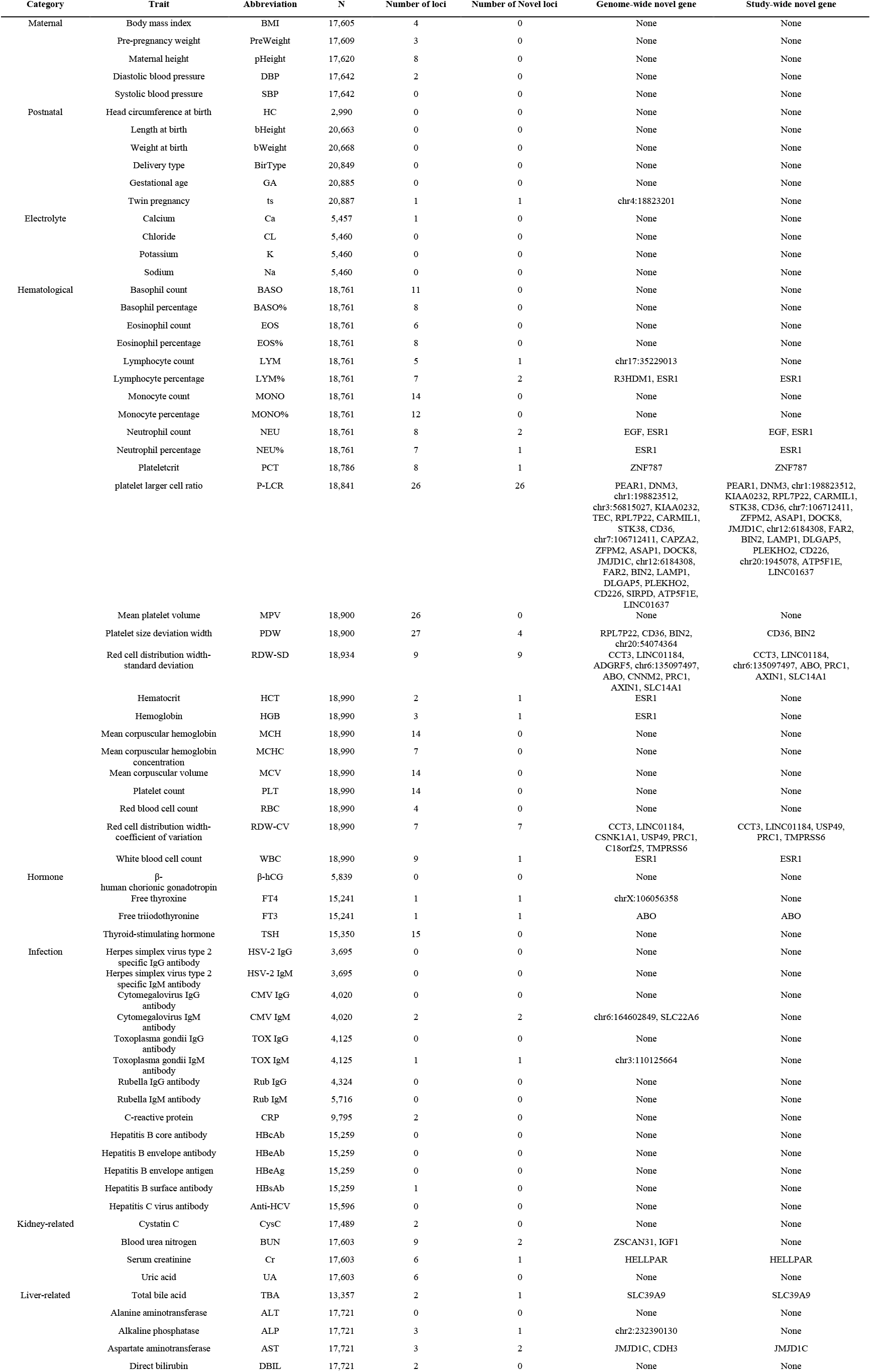
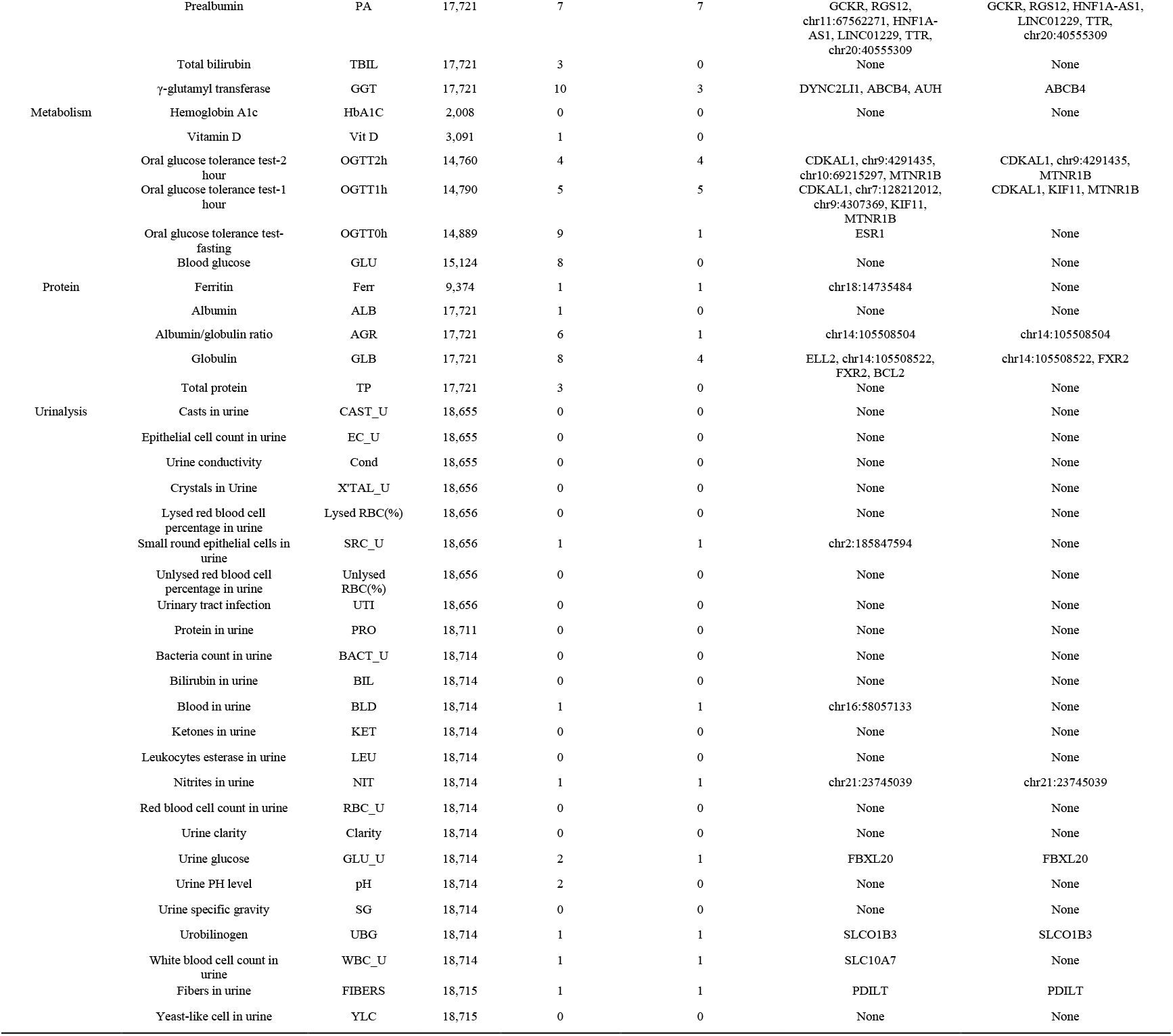
Overview of the studied pregnancy phenotypes.

At the genome-wide significance threshold of 5e-08, we identified 407 trait-locus associations involved with 66 traits (Table 1, Supplementary Table S1), the vast majority (75.18%) of which were previously reported and biologically related. For example, the fat mass- and obesity-associated (*FTO*) gene with the women’s BMI, C-reactive protein and its encoded gene *CRP*, calcium (Ca) and gene *CASR* (Calcium Sensing Receptor), the total/direct bilirubin and the members of UDP glucuronosyltransferase Family 1 (e.g., *UGT1A6*, *UGT1A8*), the vitamin D level and its associated gene *GC* (GC Vitamin D Binding Protein), etc. In addition, 101 trait-locus associations involved with 37 traits were identified for the first time, for example, the novel locus for blood urea nitrogen is *IGF1*, insulin-like growth factor 1, whose encoded protein is similar to insulin in function and structure and involved in mediating growth and development^24^.

When we considered the study-wide significance threshold of 4.81e-10 (=5e- 8/104), 272 trait-locus associations involved with 56 traits were still significant. Among them, 205 (75.37%) were previously reported and 67 associations were novel. To name a couple of novel ones: the gene *ABO* is associated with free triiodothyronine (p-value= 2.69e-31) and is previously reported to be associated with thyroid stimulating hormone measurement^25^; *FBXL20* is associated with urinary glucose and is formerly identified to play a role in kidney function^7,26^; *SLCO1B3* is associated with urobilinogen, *SLCO1B3* encodes a transmembrane receptor that plays a role in bile acid and bilirubin transport^24^; *BIN2* is associated with platelet size deviation width, previous GWAS studies have linked it with platelet count^27,28^; *JMJD1C* is associated with aspartate aminotransferase, it was identified to be associated with liver fibrosis measurement^29^, liver volume^30^, and liver enzyme^31^.

In addition, we added phenotypes currently not documented in the GWAS catalog and identified their associated variants, such as platelet larger cell ratio (P-LCR), red cell distribution width-standard deviation (RDW-SD), red cell distribution width-coefficient of variation (RDW-CV), mucus in urine, and prealbumin, etc. Among them, P-LCR had the maximum number of associated loci (n=26), including *PEAR1*, *TEC*, *STK38*, *CD36*, *JMJD1C*, *BIN2*, etc. All these genes were previously identified to be associated with platelet count and mean platelet volume^32,33^. In detail, *PEAR1* is a type of platelet receptor, *CD36* encodes a protein that is a major glycoprotein on the platelet surface and acts as a receptor for platelet-responsive proteins^24^. We identified nine loci associated with RDW-SD, including *CCT3*, *SLC12A2*, *ADGRF5*, *ABO*, *CNNM2*, *PRC1*, and *SLC14A1*, which have been previously reported to be associated with RDW^27,32^. We identified seven prealbumin-related loci, including *TTR*, *GCKR*, *HGFAC*, *HNF1A*, and *LINC01229*, in which *TTR* encodes a transthyretin protein, a type of prealbumin, and transports thyroid hormones in plasma and cerebrospinal fluid, and *HNF1A* encodes a protein that is a transcription factor required for the expression of several liver-specific genes and an albumin proximal factor^24^. Mucus in urine is associated with *UMOD* (p-value = 1.28e-17), which was previously identified to be associated with urinary uromodulin in the European population^34^ and its encoded protein was most abundant in mammalian urine.

We also discovered some potentially pregnancy-specific associations. In detail, *ESR1* with fasting serum glucose, hemoglobin, hematocrit, and several types of leukocytosis (white blood cell, neutrophils, lymphocytes). The protein encoded by *ESR1* (Estrogen Receptor 1) regulates the transcription of many genes that play a role in gestation, metabolism, sexual development, growth, and other reproductive functions^24^. We suspect that, during the special pregnancy period, significant changes in hormonal levels have highlighted the associations between *ESR1* and pregnancy phenotypes. For blood urea nitrogen, we found a novel gene *ZSCAN31*, which encodes a protein containing multiple zinc finger motifs and may be involved in the development of multiple embryonic organs^35^. We also identified the novel association between *ABCB4* with γ-glutamyl transferase. The protein encoded by *ABCB4* is a member of the superfamily of ATP-binding cassette transporters and may involve the transport of phospholipids from the liver into bile. A previous study demonstrated that the splicing mutations in *ABCB4* can cause intrahepatic cholestasis of pregnancy in women with high γ-glutamyl transferase^36^.

To detect the pleiotropy effects of the significant loci, we counted the number of their identified times (Supplementary Table S2), and a few interesting findings were observed. Among all associations, the loci *MED24*/*PSMD3*/*CSF3/THRA* (CHR17: 39,980,807-40,093,867) were identified the most. The associated phenotypes all belong to the leukocyte group, such as neutrophils, eosinophils, and basophils. The following genes are *ESR1*, *ABO*, and *JMJD1C*. Among them, *ESR1* was identified to be associated with fasting serum glucose, white blood cell counts, neutrophils counts and ratio, lymphocytes ratio, hemoglobin, and hematocrit; all *ESR1*-trait associations were novel findings. The gene *ABO* was associated with alkaline phosphatase, mean corpuscular hemoglobin concentration, mean corpuscular volume, thyroid-stimulating hormone, free triiodothyronine, and red blood cell distribution width-standard deviation; the former four associations were known, while the latter two were novel. For gene *JMJD1C*, the known associated phenotypes included γ-glutamyl transferase, platelet count, platelet size deviation width, and mean platelet volume; while newly identified associated traits were platelet larger cell ratio and aspartate aminotransferase. We used the phenome-wide association study (PheWAS) Manhattan plots to show the associations between one genetic variant and different phenotypes (Figure 3).

**Figure 3.**
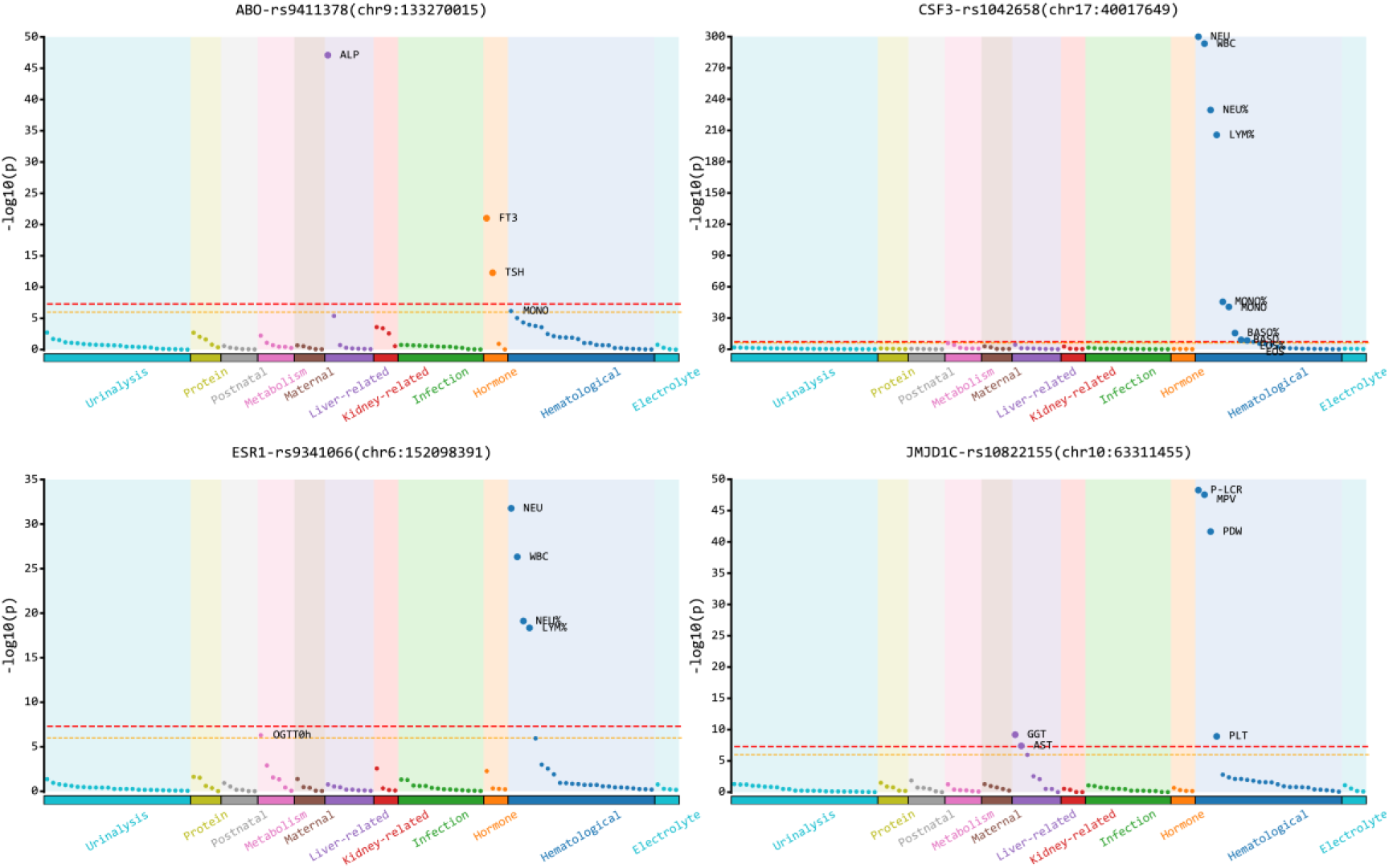
PheWAS Manhattan plots. *Notes*: the GWAS associations for four listed SNPs and phenotypes in different categories. The yellow dashed line indicates -log10(1e-06) and the red dashed line indicates -log10(5e-08).

The number of identified loci and calculated heritability for each phenotype were provided in Figure 2c. The heritability analysis showed that there were 51 phenotypes with heritability of 10% or more, with SE less than 5% in 36 of them. The results were quite consistent with that of GWAS and most of the phenotypes with high heritability belonged to hematological, followed by liver-related, kidney-related, infection, and the lowest was urinalysis. Among the phenotypes with SE less than 5%, platelet larger cell ratio (P-LCR) had the largest heritability of 32.0%. The heritability of maternal height and BMI were 30.0% and 26.8%, respectively, which were 26.0% and 0.2% less than previously reported results^37,38^. The main reason is that the coverage of ultra-low depth sequencing data is only 0.6%-1% of the entire genome, resulting in many undetected variants. Even after high-quality genotype imputation, only about 2 million SNPs were included in the analysis after the quality control, thus the explained variation of the phenotype is lower than that of high-depth sequencing data.

### Pathway enrichment analysis

The detailed results for all phenotypes were provided in Supplementary Table S3. We also listed the most significant pathway for each phenotype in Figure 4. At an empirical p-value less than 1e-04, 24 phenotypes had at least one significant pathway. Among them, the lymphocyte count had the maximum number of 14 associated pathways, followed by eosinophil percentage, eosinophil count, and γ-glutamyl transferase.

**Figure 4.**
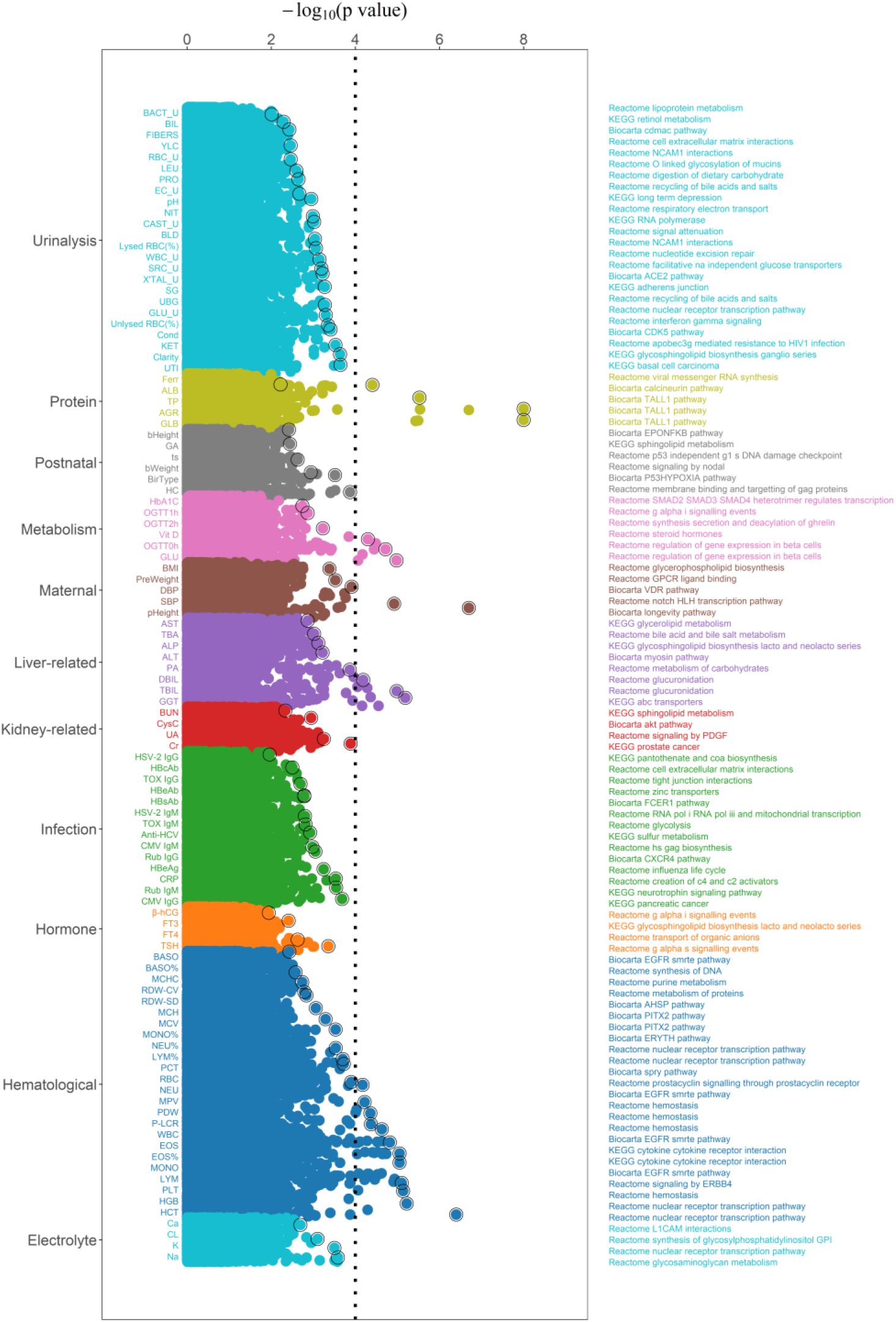
The gene-set enrichment analysis based on SNP-based GWAS results. *Notes*: The x-axis represents the -log10 transformed empirical p-values for pathways. The most significantly associated pathway with each trait was labeled.

We found that the most important associated pathways belonging to the same category were often the same or similar. For example, pathways associated with the number and percentage of leukocytes included inflammatory molecular signaling pathways (e.g., BioCarta IL17 pathway), cytokine pathways involved in adaptive inflammatory host defense and cell growth (e.g., KEGG cytokine-cytokine receptor interaction), and epidermal growth factor receptor gene (*EGFR*) involved in pathways associated with a variety of human diseases (e.g., BioCarta EGFR SMRTE pathway). For glucose levels, such as serum glucose and oral glucose tolerance test measurements, the associated pathways were related to the development of pancreatic islet B cells (Reactome regulation of beta cell development), type II diabetes (KEGG maturity-onset diabetes of the young), and hormones (Reactome peptide hormone biosynthesis), all of which played important roles in glucose metabolism. Notable pathways associated with globulin, albumin ratio, and total protein are mainly immune-related pathways, such as BioCarta TALL1 pathway, KEGG intestinal immune network for IGA production, and KEGG primary immunodeficiency.

In addition, for a single phenotype, the most relevant pathway for bile acids is Reactome bile acid and bile salt metabolism, which mainly describes the synthesis and metabolism of bile acids and bile salts; the most significant pathway for platelet counts is Reactome hemostasis, which describes the physiological response of the body to hemostasis, including vasoconstriction, platelet thrombosis, and fibrin clot formation.

The enriched pathway for total and direct bilirubin levels are Reactome glucuronidation and KEGG pentose and glucuronate interconversions, which are involved in bilirubin metabolism. For fasting glucose levels in OGTT test, the enriched pathway is Reactome regulation of gene expression in beta cells, which is associated with beta cells and insulin synthesis.

Besides these previously known functional pathways, we also uncovered several candidate ones that might act in performing biological functions in phenotypes. The most significant pathway for birthweight is Reactome signaling by NODAL. The *NODAL* gene encodes a TGF-beta (transforming growth factor-beta) superfamily of proteins, which regulates early embryonic development and plays important roles in the maintenance of human embryonic stem cell pluripotency and placental development. The related pathways for maternal height included BioCarta GH pathway, Reactome growth hormone receptor signaling, and Reactome prolactin receptor signaling. The former two pathways were about growth hormones and played a major role in regulating growth during childhood and adolescence. A deficiency in growth hormone signaling can cause dwarfism. Interestingly, we found that prolactin receptor signaling also achieved molecular function in maternal height. Prolactin is a hormone secreted mainly by the anterior pituitary gland and regulates the development of the mammary gland and lactation. Whether this pathway plays a role specifically in female height requires further investigation. Among the few top functional pathways related to a twin pregnancy, we observed Reactome cell cycle checkpoints and Reactome mitotic G1- G1/S phases. The formation process of identical twins is mitotic, and we speculate that these two pathways play an important role in twin pregnancy.

### Partitioning heritability analysis

The majority of the partitioning heritability results recapitulate our known biology knowledge (Figure 5, Supplementary Figure S5, Supplementary Table S4): leukocyte phenotypes (basophils, eosinophils, lymphocytes, monocytes) exhibit blood/immune cell-type enrichments; liver-related phenotypes (aspartate aminotransferase, total bilirubin, direct bilirubin, alkaline phosphatase, alanine aminotransferase) exhibit, liver, CNS, and blood/immune enrichments; virus infection-related phenotypes (HBsAb, HBcAb, HBeAb, HBeAg, Anti-HCV) exhibit blood/immune cell-type enrichments. For a particular phenotype, maternal height exhibits musculoskeletal/connective enrichment; leukocyte esterase in urine exhibits blood/immune cell-type enrichment; urine clarity exhibits bladder enrichment; blood urea nitrogen exhibits urinary bladder cell-type enrichment; thyroid-stimulating hormone exhibits thyroid enrichment.

**Figure 5.**
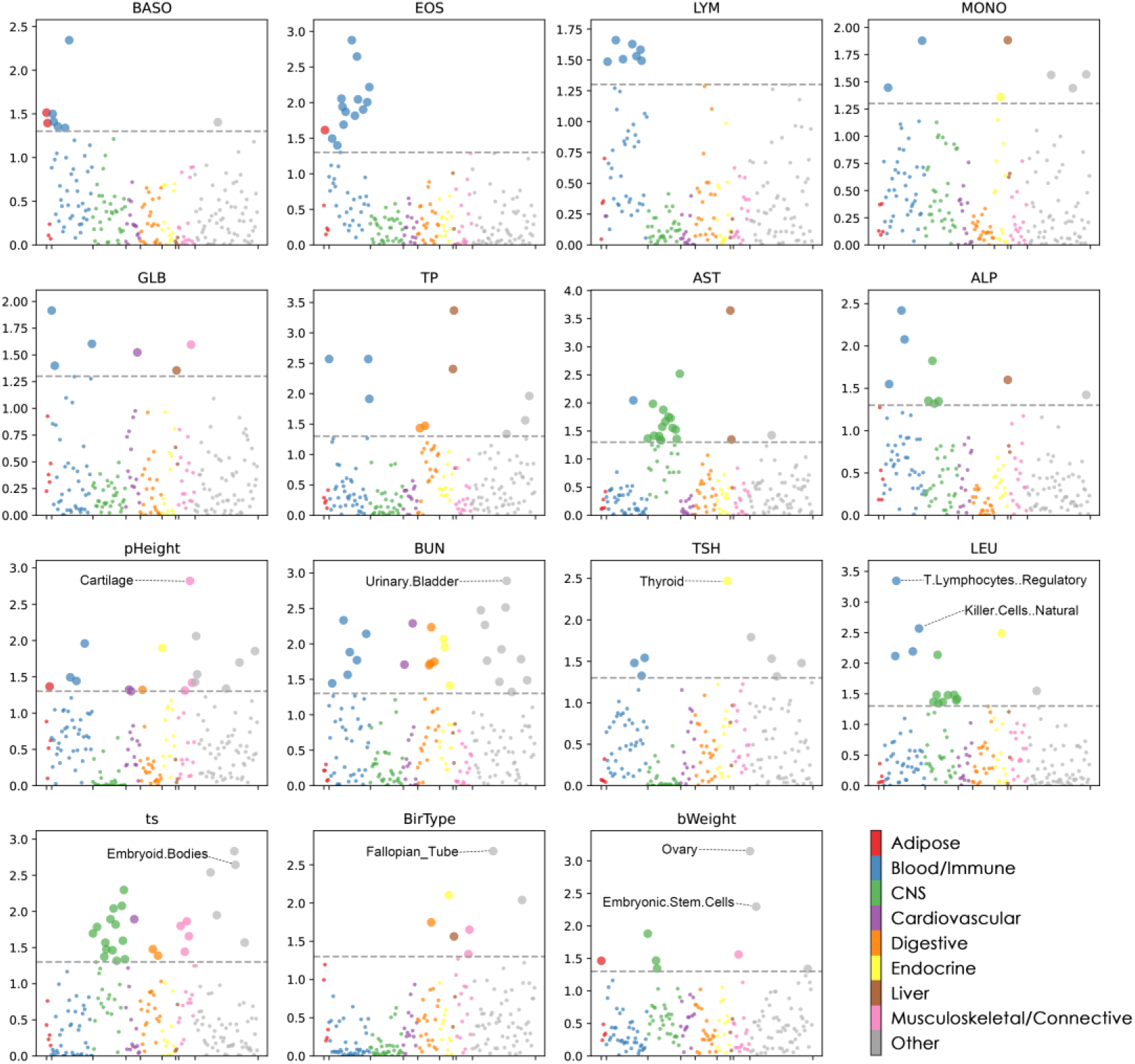
Results of the partitioning heritability analysis for selected traits. *Notes*: Each point represents a tissue/cell type from either the GTEx data set or the Franke lab data set. Large points pass the p-value < 5% cutoff, –log10(P)=1.30.

In addition to the aforementioned trait-tissue/cell-type relevance, we also noticed a few other interesting ones (Figure 5, Supplementary Table S4), which were closely related to pregnancy. The twin pregnancy exhibits embryoid bodies cell-type enrichment. The birth-type cesarean section exhibits fallopian tube cell-type enrichment. The birth weight and birth height exhibited ovary and embryonic stem cell enrichments. Previous research reported that birthweight was a relation to maternal ovarian size^39,40^.

### Replication in GWAS of pregnancy phenotypes

As a replication strategy, we compared our GWAS results with three companion works to discover shared signals [cite]. For simplicity, we denote our work as PP (pregnancy phenotype), and the companion works are MM (maternal metabolite), NM (neonatal metabolite), and EHR (electronic health record). In total, we found 210 shared significant SNPs that were composed of 18 loci linking 19 representative pairs of possibly correlated traits (Supplementary Table S5). Some of the findings are well-established, such as PP:Vitamin_D─MM:Vitamin_D3 with *GC* gene (PP4 = 99.9%)^41–43^, PP:MCH─MM: Element_Fe with *TMPRSS6* gene (PP4 = 100%)^11,44,45^, and PP:BMI─EHR:Obesity with *FTO* gene (PP4 = 96.7%)^46–49^, where MCH is mean corpuscular hemoglobin. Here, PP4 indicates posterior probability of H4: one common causal variant of two GWAS studies. Some discoveries are novel but convincing, such as PP:Prealbumin─MM:Vitamin_A with *GCKR* gene (PP4 = 99.5%), PP:CR─MM:Vitamin_A with *GCKR* (PP4 = 98.6%), PP:MCH─NM:C2 with *MARCH8* gene (PP4 = 98.9%), PP:CR─MM:Element_Mg with *CASP9* gene (PP4 = 95.6%), PP:UA─MM:Element_Mg with *DNAJC16* gene (PP4 = 99.2%), and PP:FT4─MM:Element_I with *SERPINA7/PWWP3B* (nearest) gene in X chromosome (PP4 = 94.3%), where CR is serum creatinine, C2 is acetylcarnitine, UA is urine acid, and FT4 is free thyroxine. Specifically, prealbumin was initially discovered to function as a transport protein for thyroxine and vitamin A, and an intronic variant (rs780094) on *GCKR* was found as a significant positive association with vitamin A^50^, suggesting the likely association of prealbumin and *GCKR*. An early study reported that in the US population, vitamin A and CR were positively correlated with each other^51^, while CR was associated with *GCKR*^11,52^. Several studies have shown that higher magnesium levels were associated with lower risk of hyperuricaemia^53,54^, while urine acid had significant gene *DNAJC16*^27,55^. The plots for visualizing genetic colocalization analysis of two GWAS results from this study and the companion works were provided in Figure 6 and Supplementary Figure S6. The replication study validated our GWAS findings.

**Figure 6.**
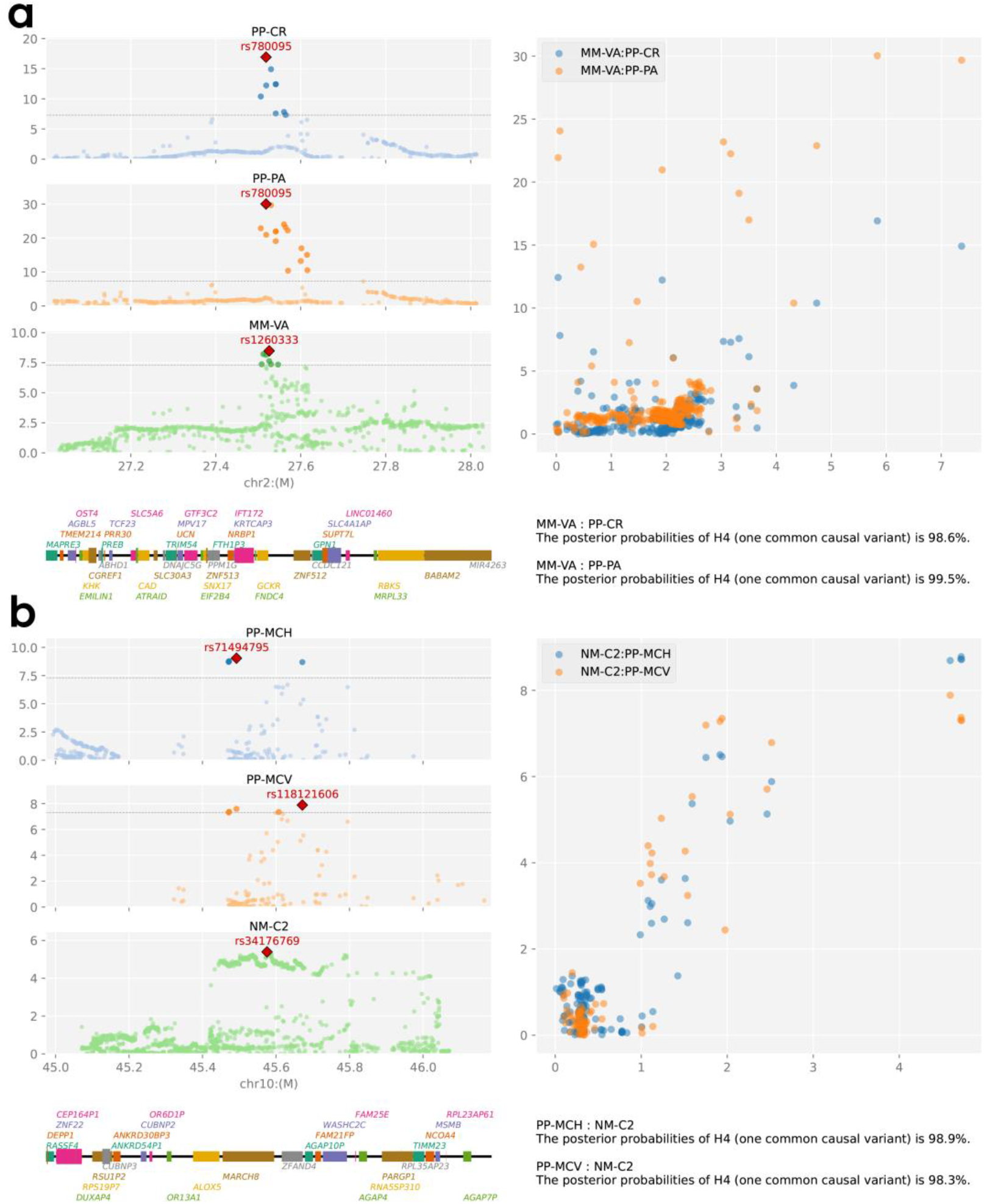
The genetic colocalization analysis of our study and the companion works. *Notes*: **a-b)** plots for visualizing colocalization analysis for pairs of phenotypes from this study and the companion works, where PP is pregnancy phenotypes, MM is maternal metabolites, NM is neonatal metabolites, CR is serum creatinine, prealbumin, VA is vitamin A, MCH is mean corpuscular hemoglobin, MCV is mean corpuscular volume, and C2 is acetylcarnitine.

## Methods

### Study subjects

All the subjects were recruited from Wuhan Children’s Hospital, Wuhan city, Hubei province, China during their pregnancy routine tests between 2015 to 2020. Wuhan is the most populous city in Central China. This study was approved by the Institutional Review Boards at Wuhan Children’s Hospital and BGI-Shenzhen and also approved by the Human Genetic Resources Administration of China.

### Genotype and imputation

The genotype data were generated when pregnant women underwent NIPT screening from around the 12^th^ week of pregnancy onwards. In detail, 5ml peripheral venous blood was collected and stored at -80 Celsius degrees in EDTA-anticoagulated tubes. Plasma samples from pregnant women were sent to BGI-Wuhan for next-generation high-throughput sequencing, and genotype data with an ultra-low sequencing depth of 0.06X to 0.1X were obtained for each subject. FASTP software was used to remove read regions of low quality and potential adaptor sequences^56^. For genome alignment, single-end reads (35bp) were aligned to the human reference genome (GRCh38/hg38) using the BWA algorithm^57^. Then, we used the BaseVar algorithm to call SNPs and STITCH to impute the missing genotypes^58^. Finally, we filtered out samples if the sequencing depth was lower than 0.05X or the mapping rate was less than 90%.

To evaluate the genotype imputation performance, we used 30 randomly selected Han Chinese from the 1000 Genomes Project with 30X sequencing coverage (downloaded from https://www.internationalgenome.org/data-portal/data-collection/30x-grch38) as a true set, down-sampled to 0.1X sequencing depth, and imputed with the true NIPT genotype data by using STITCH. The imputation accuracy was computed as Pearson’s correlation coefficient between the 30X true set and the imputed genotype. In addition to this assessment, we evaluated the imputation performance by comparing the imputed and true genotypes of three variants in two genes (*MTHFR* and *MTRR*), which were involved in the folic acid metabolism. In the obstetrical clinic, these three variants were often tested for folate metabolism ability in pregnant women. Mutation carriers in these variants may have a higher risk of miscarriages or birth defects^59^.

### Phenotype

A wide spectrum of traits was measured during the pregnancy routine tests, including basic maternal information, postnatal outcome, and laboratory measurements. The basic maternal information includes pregnant women’s age, height, weight, and blood pressure. The postnatal outcome includes gestational week, delivery method, birth weight, and birth length. The laboratory measurements include various serum and urinary laboratory tests, such as hematology tests, liver function, urine sediment analysis, and viral infections. Since the body will undergo some adaptive or pathological changes as pregnancy progresses, pregnant women need to take some examinations more than once and see if the relevant indicators are normal. Multiple examinations during the entire pregnancy period produced multiple records. For quantitative traits, we took an average of multiple measurements, and for binary traits, we treated as positive once a positive result was observed. Detailed characteristics for each trait are shown in Table 1.

### Genome-wide association analyses

For each quantitative trait, we performed a GWAS analysis by fitting a linear regression model; and for a binary trait, we fitted a logistic model. This was achieved in PLINK 2.0 using the command line –*glm*^60^. The imputed genotype was measured in a dosage ranging from 0 to 2. The biallelic variants with minor allele frequency (MAF) > 0.05, Hardy-Weinberg equilibrium (HWE) p-value > 1e-6, and genotype missing rate < 0.1 were used in the GWAS analyses. The covariates adjusted in the model include women’s age and the first five principal components (PCs). We set a genome-wide significance threshold of 5e-08 and a study-wide significance threshold of 4.81e-10 (=5e-8/104) by performing Bonferroni adjustment.

We defined an associated locus with a window size of 1MB based on its physical position in the genome and counted the number of loci associated with the phenotype. For a locus, if all SNPs it contains were less than 500 kb away from the reported trait-associated SNPs in the GWAS catalog as of September 2022 (https://www.ebi.ac.uk/gwas/), we defined this locus as known, and vice versa as novel.

The genomic inflation factors (λgc) were then calculated in R^61^. With the GWAS summary statistics of each trait, we used LD score regression (LDSC)^62^ to estimate its heritability and confounding bias.

### Pathway enrichment analysis

We used Pascal to calculate gene and pathway scores on summary data from GWAS^63^. In particular, Pascal used the reference population to calculate LD information, and the reference population used in this case was the Asian population (1000 genome phase 3 Asian population). Pascal used three external databases to define the gene set of each pathway, including BIOCARTA^64^, KEGG (Kyoto Encyclopedia of Genes and Genomes)^65^, and REACTOME^66^, with 1,077 gene sets. The window size set for this experiment was 50kb, and the significance of the path was evaluated using the empirical score. We used 1e-4 as the significance threshold.

### Partitioning heritability analysis

We applied stratified LD score regression^67^ to estimate the polygenic contributions of functional categories to heritability in each trait. We used 205 cell-type-specific annotations with gene expression data from the Genotype-Tissue Expression (GTEx) project^68^ and Franke lab dataset^69,70^. The 205 tissues and cell types were classified into nine categories, including adipose, blood/immune, cardiovascular, central nervous system (CNS), digestive, endocrine, liver, musculoskeletal/connective, and others. This classification is referenced to Finucane et al.^71^. The multi-tissue microarray gene expression file is “Multi_tissue_gene_expr.EAS” containing both the GTEx dataset and the Franke lab dataset with East Asian populations. The baseline model LD score was 1000G_Phase3_EAS.

### Replication study of GWAS candidate loci

Ideally, the replication study of GWAS should be re-running the association inference conducted on independent samples from the same population, while the sample size should be as large as or larger than the discovery sample. However, an independent dataset or GWAS summary statistics of the studied pregnancy phenotypes are largely not accessible in Chines or East Asian women. As an alternative to this, we compared the identified SNP-trait associations in three companion works, which performed GWAS analysis for maternal metabolites [cite], neonatal metabolites [cite], and electronic health records for both pregnant women and newborns [cite], respectively. Specifically, we searched for shared genome-wide significant SNPs between our work and each of the external work. If any, we defined a genomic region of 1-Mb with 500-kb on each side of the lead SNP and performed colocalization analysis in *R:coloc* to test whether the two traits shared distinct or same causal variants^72^.

## Discussion

Pregnancy is a special period experienced by women, in which regular maternal examinations generate a large amount of systematic clinical data, however, there are few genetic studies targeting these pregnancy phenotypes. In recent years, with the popularity of noninvasive prenatal genetic testing technology, it has become possible to obtain high-throughput maternal genotype data. In this study, based on the genotype data of nearly 40,000 pregnant women with non-invasive prenatal genetic testing, we performed molecular biology analysis of hundreds of clinical maternal phenotypes and successfully replicated 75.18% of the trait-locus associations and also mined 101 novel loci involved with 37 phenotypes at the genome-wide significance threshold; the gene-set enrichment analysis not only further confirmed the phenotype-related pathways of action, but also discovered several pregnancy-specific associations; the results of heritability and partitioning heritability quantified the influence of genetic factors and highlighted valuable relevant tissue/cell types functioning in each phenotype.

Although our study provided an important reference and data resource for studying pregnancy phenotypes and complications, there are still several limitations. First, even though the genotype dataset was available in nearly 40,000 samples and the pregnancy traits were collected from over 30,000 women, the effective sample size in GWAS analysis was only around 20,000 after matching the genotype and phenotype datasets. The GWAS analysis requires larger sample sizes to achieve sufficient statistical power. Thus, we aim to collect more samples for performing larger-scale genetic studies in the future. Second, our phenotypes were all limited to clinical information and laboratory test indicators, and there is a lack of maternal disorders and adverse postnatal outcomes. Obtaining information on diseases diagnosed in the electronic medical record (EMR) system for pregnant women and analyzing them with pregnancy laboratory indicators such as genetic correlation^73^ and causal inference are our further research directions.

As the growing of NIPT sequencing data, we expect more and larger genetic studies on maternal- and neonatal-related traits and diseases, the genome-wide association results would provide valuable reference and shed light on maternal and child health care. A systematic cataloguing and summarization of these associations would be highly effective for researchers to find what they need. Several websites have been developed as a collection of GWAS summary datasets studied on a wide variety of phenotypes and also available for researchers to browse and download, for example, the GWAS Catalog (https://www.ebi.ac.uk/gwas/home), the MRC Integrative Epidemiology Unit (IEU; https://gwas.mrcieu.ac.uk/), the PheWeb archive (https://pheweb.sph.umich.edu/), and the BioBank Japan PheWeb (https://pheweb.jp/). To offer a searchable, visualizable, and openly accessible database of pregnancy-related SNP-trait associations, we are currently building a free online website for sharing and visualizing the GWAS results reported by this work, study of maternal metabolites [cite], neonatal metabolites [cite], and EHR of maternal and neonatal [cite]. We anticipate that the website could play a fundamental role in studying genetic susceptibility of maternal/postnatal-related phenotypes and providing useful resources for scientists, clinicians, and other users worldwide.

## Author contribution

X.J., A.Z., and H.Z. conceived the study, designed the research program, and managed the project.

H.X., M.Y., J.Z., Y.Z., J.L., Y.Y.Z., Z.C., H.M., X.C., L.H., and R.Z. collected the data.

L.L., Y.T., Y.H., and P.L. preprocessed the data and finished the quality control.

H.Z., H.X., L.L., M.Y., J.Y.Z., M.C., and P.L. performed the statistical analysis and results visualization.

H.Z., H.X., L.L., M.Y., and M.C wrote the manuscript. All authors participated in revising the manuscript.

## Acknowledgment

This study was supported by the Central Guidance on Local Science and Technology Development Fund of Hubei Province (2022BGE261), National Natural Science Foundation of China (32171441 and 32000398), Guangdong-Hong Kong Joint Laboratory on Immunological and Genetic Kidney Diseases (2019B121205005), Top Medical Young Talents (2019) of Hubei Province, Guangdong Provincial Key Laboratory of Genome Read and Write (2017B030301011), Open project of BGI-Shenzhen, Shenzhen 518000 China (BGIRSZ20200008), and the China National GeneBank.

## Data availability

The GWAS summary statistics of all studied phenotypes have been deposited into CNGB Sequence Archive (CNSA)^74^ of China National GeneBank DataBase (CNGBdb)^75^ with accession number CNP0003673.

## Supplementary Figures

**Supplementary Figure S1**. The number of available samples for each trait

**Supplementary Figure S2**. The visualization of trait distribution and pairwise association

**Supplementary Figure S3**. The Manhattan plots of all studied traits

**Supplementary Figure S4**. The heatmap of partitioning heritability results

**Supplementary Figure S5**. The results of partitioning heritability for traits that were not listed in the main text

**Supplementary Figure S6**. The genetic colocalization analysis of trait pairs that were not listed in the main text

